# Improved prediction of the response duration to MAPK inhibitors in patients with advanced melanoma using baseline genomic data and machine learning algorithms

**DOI:** 10.1101/2023.12.07.23299389

**Authors:** Sarah Dandou, Kriti Amin, Véronique D’Hondt, Jérôme Solassol, Olivier Dereure, Peter J. Coopman, Ovidiu Radulescu, Holger Fröhlich, Romain M. Larive

## Abstract

**Purpose:** Baseline genomic data have not demonstrated significant value for predicting the response to MAPK inhibitors (MAPKi) in patients with BRAF^V600^-mutated melanoma. We used machine learning algorithms and pre-processed genomic data to test whether they could contain useful information to improve the progression-free survival (PFS) prediction.

**Experimental design:** This exploratory analysis compared the predictive performance of a dataset that contained clinical features alone and supplemented with baseline genomic data. Whole and partial exon sequencing data from four cohorts of patients with BRAF^V600^-mutated melanoma treated with MAPKi were used: two cohorts as training/evaluation set (n = 111) and two as validation set (n = 73). Genomic data were pre-processed using three strategies to generate eight different genomic datasets. Several machine learning algorithms and one statistical algorithm were employed to predict PFS. The performance of these survival models was assessed using the concordance index, time-dependent receiver operating characteristic (ROC) curve and Brier score.

**Results:** The cross-validated model performance improved when pre-processed genomic data, such as mutation rates, were added to the clinical features. In the validation dataset, the best model with genomic data outperformed the best model with clinical features alone. The trend towards improved prediction with baseline genomic data was maintained when data were censored according to the two clinical setting scenarios (duration of clinical benefit and progression before 12 months).

**Conclusion:** In our models, baseline genomic data improved the prediction of response duration and could be incorporated into the development of predictive models of response pattern to MAPKi in melanoma.

**Translational Relevance:** Currently, biomarkers are lacking to robustly predict the response to therapy targeting the MAPK pathway in advanced melanoma. Therefore, in the clinic, a trial-and-error approach is often used. Baseline genomic mutation profiles represent a comparably stable biological readout that is easily accessible and measurable in clinical routine. Therefore, they might represent candidate predictive biomarker signatures. However, previous studies could not show a clear predictive signal for the response to MAPK inhibitors (MAPKi) in patients with BRAF^V600^-mutated melanoma. Here, our exploratory machine learning-based analysis highlighted an improved prediction of progression-free survival when clinical and genomic data were combined, even when using only partial exome sequencing data. This suggests that baseline genomic data could be incorporated in the development of predictive models of the response to MAPKi in advanced melanoma by leveraging the results of current routine partial exome sequencing.

**Interest statement:** The authors declare no potential conflicts of interest.

## 1 Introduction

MAPK inhibitors (MAPKi) and immune checkpoint inhibitors (ICI) have totally revolutionized the treatment of patients with advanced melanoma [1, 2, 3].However, their efficacy is hampered by primary or secondary tumor resistance. Patients with melanoma harboring BRAF^V600^ mutations show a good overall response rate to MAPKi at treatment start, while ICI have a more durable response. Therefore, the choice between these therapeutic options requires a robust prediction of the response and its duration.

Baseline genomic data represent a reliable and easily accessible source of information to identify predictive biomarkers. However, for MAPKi response prediction, previous studies failed to show any predictive value of baseline genomic data [4, 5], with the exception of TERT promoter mutations [6, 7]. Conversely, tumor mutation burden is an U.S. Food and Drug Administration-approved biomarker of the response to ICI [8]. Therefore, research on biomarkers of MAPKi response has progressively moved from baseline tumor genomic data to melanoma cell identity [9], tumor microenvironment immune components [10] and blood proteins [11]. Machine learning models have already shown their value in prognosis and diagnosis [12]. Recently, machine learning approaches have been developed to identify predictive biomarkers of the response to ICI [11, 13, 14, 15], and also to predict early-stage melanoma recurrence [16, 17] and sentinel lymph node status [18].

In this study, we employed several machine learning models to assess the value of baseline genomic data to predict MAPKi response. To investigate a relevant number of patients, we collected four independent cohorts of patients with melanoma treated with MAPKi and with pre-treatment whole or partial tumor exome sequencing data. Our results showed a significant improvement in the accuracy of progression-free survival (PFS) prediction when pre-processed mutation data were added to the clinical characteristics of the two cohorts used for the training/evaluation step. We validated this observation using the other two cohorts, and confirmed the prediction improvement in two clinical setting scenarios (duration of clinical benefit and disease progression before 12 months).

## 2 Materials and Methods

### 2.1 Patient data and Tumor specimens

We collected the data of four cohorts of patients with BRAF^V600E/K^-mutated melanoma treated with MAPKi: (1) Blateau et al. (vemurafenib, vemurafenib and cobimetinib, dabrafenib, dabrafenib and trametinib; *n*=53) [6]; (2) Catalanotti et al (vemurafenib, vemurafenib and cobimetinib, dabrafenib, dabrafenib and trametinib; *n*=66) [19]; (3) Louveau et al. (vemurafenib and cobimetinib, dabrafenib and trametinib; *n*=20) [5]; (4) Van Allen et al. (vemurafenib, dabrafenib; *n*=45) [20]. Pretreatment tumors were from patients with stage IV or unresectable stage III BRAFV600E/K melanoma. Clinical responses to therapy were determined using RECIST criteria [21]. The use of data from these patient cohorts has been approved by the ethics committee of the University of Montpellier, France (favorable advice UM 2022-009bis). As we did not generate new data for this study, no additional ethics approval was required. For each patient, tumor tissues were collected, stored as formalin-fixed, paraffin-embedded (FFPE), or frozen samples, and processed. Somatic single-nucleotide variants and small insertions or deletions were considered as mutations. Table 1 summarizes the patients’ demographic/clinical features, outcomes, and DNA sequencing methods/results.

**Table 1:** Patients, treatment outcomes and genomic data.

| Feature | Values | Total (%) | Van Allen et al. | Blateau et al. | Catalanotti et al. | Louveau et al. |
| --- | --- | --- | --- | --- | --- | --- |
| Total number of patients | - | 184 | 45 | 53 | 66 | 20 |
| Sex | male<br>female | 101 (55 %)<br>83 (45 %) | 22 (48 %)<br>23 (52 %) | 28 (53 %)<br>25 (47 %) | 41 (62 %)<br>25 (38 %) | 10 (50 %)<br>10 (50 %) |
| Age | Median (range), years | 54 (18-94) | 51 (25-76) | 57 (21-90) | 54.5 (21-83) | 51.5 (18-94) |
| Lactate dehydrogenase | elevated<br>normal<br>missing | 53 (29 %)<br>63 (34 %)<br>68 (37 %) | 6 (13 %)<br>-<br>39 (87 %) | 23 (43.3 %)<br>25 (47 %)<br>5 (9.43 %) | 24 (36 %)<br>38 (57.5 %)<br>4 (6 %) | -<br>-<br>20 |
| Type of drug | vemurafenib<br>dabrafenib + trametinib<br>dabrafenib<br>vemurafenib + cobimetinib<br>N.A | 83 (45%)<br>67 (4%)<br>22 (12%)<br>11 (6%)<br>1 (0.5%) | 31 (69 %)<br>-<br>14 (31 %)<br>-<br>- | 4 (7.5 %)<br>42 (79 %)<br>2 (3.7 %)<br>4 (7.5 %)<br>1 (2 %) | 48 (72 %)<br>8 (12 %)<br>6 (9 %)<br>4 (7 %)<br>- | -<br>17 (85 %)<br>-<br>3 (15 %)<br>- |
| Best Overall Response | progressive disease<br>stable disease<br>partial response<br>complete response<br>N.E = not evaluated | 59 (32 %)<br>30 (16 %)<br>70 (38 %)<br>19 (10 %)<br>4 (2 %) | 7 (15 %)<br>12 (27 %)<br>25 (56 %)<br>1 (2 %)<br>- | 40 (78 %)<br>3 (6 %)<br>5 (9 %)<br>3 (6 %)<br>2 (1 %) | 10 (15 %)<br>13 (20 %)<br>31 (47 %)<br>8 (12 %)<br>4 (6 %) | 2 (10%)<br>2 (10%)<br>9 (45 %)<br>7 (35 %)<br>- |
| Progression-free survival - months | 0.7-68 (median=6.0) | median | median=5.2 | median=6.1 | median=5.75 | median=9.5 |
| Overall survival - months | 0.7-69 (median=13.5) | median | - | median=10.5 | median=14.5 | median=28 |
| Vital status | 1=dead<br>0=alive | 90 (65 %)<br>48 (34 %) | -<br>- | 37 (71 %)<br>15 (29 %) | 45 (68 %)<br>21 (31 %) | 8 (40 %)<br>12 (60 %) |
| Sequencing method | Whole exome or targeted sequencing | - | WES | Targeted (35 genes) | Targeted (300 genes) | Targeted (74 genes) |
| Mutated genes (%) |  |  | 5.32 % | 5 % | 18.8 % | 2 % |

Summary of clinical features, treatment outcomes and genomic data collected for the study. Each outcome is detailed by cohort. Categorical outcomes are described with numbers and percentages, and continuous outcomes using the median of the distribution for each cohort. The number of sequenced genes and the percentage of mutated genes in each cohort are indicated.

### 2.2 Genomic data pre-processing

#### 2.2.1 Functional consequences of mutations detected in tumor samples

We used FATHMM [22], a software that predict the functional consequences of the coding and non-coding variants detected by genome sequencing. We used a binary classification to categorize genes in function of the FATHMM results. If a somatic mutation detected in a gene was predicted to be a cancer-promoting/driver mutation by FATHMM, that gene was classified in column 1, otherwise in column 0. We then selected the genes mutated in at least 5% of the patients (*Fathmm* dataset). Alternatively, we used Cscape [23], that predicts the oncogenic status (i.e. disease-driver or neutral) of somatic point mutations in the coding and non-coding regions of the cancer genome. As done with FATHMM, we employed the obtained information to define a feature set consisting of binary indicators and selected the genes mutated in at least 5% of the patients (*Cscape* dataset). We also defined variants for which only high confidence predictions by Cscape were considered (*Cscape*^*High*^ dataset).

#### 2.2.2 Selection of genes involved in melanoma signaling pathways

To select genes involved in melanoma signaling pathways, we used the curated melanoma pathway from the KEGG database [24] and the curated melanoma pathway model from the Virtual Melanoma Cell (VCELLS) project [25]. We only considered the 70 proteins of the KEGG melanoma pathway (*KEGG-Mel* dataset), and the 119 proteins of the VCELLS melanoma pathway (*VCELLS-Mel* dataset), both represented by HGNC gene symbols.

#### 2.2.3 Pathway enrichment analysis

As an alternative to the previously described binary indicator approach, we constructed features by statistical overrepresentation analysis (hyper-geometric test) of mutated genes in all KEGG pathways (*MR*^*KEGG*^ dataset) and in the KEGG melanoma (*MR*^*KEGG-Mel*^ dataset) and VCELLS melanoma (*MR*^*VCELLS-Mel*^ dataset) pathways [26]. We performed this analysis for each patient separately, and obtained one p-value per patient and per pathway. Because p-values span over several decades, we used the negative logarithm of this p-value as a feature value.

### 2.3 Survival analysis algorithms

#### 2.3.1 Elastic net penalized Cox regression (EN-COX)

Penalized models introduce regularization by adding penalty terms to the model equation. The Elastic Net penalty regularization method is particularly useful in scenarios with more features than observations. It combines the strengths of lasso (*l*_1_) and ridge (*l*_2_) penalties, by performing feature selection like lasso and by shrinking correlated features together like done in ridge regression [27].

#### 2.3.2 Random survival forest (RSF)

RSF is an ensemble method for survival analysis that uses survival trees as its base model. Unlike the decision trees used in Random Forests (described by Breiman), RSF employs survival trees and averages the ensemble cumulative hazard function of each tree for prediction [28].

#### 2.3.3 Extra survival trees (EST)

EST, also known as extremely random survival forest, is a meta-estimator developed as an extension of the RSF method. This method fits several randomized survival trees, called extra-trees, and like RSF, uses averaging to improve the predictive accuracy and control overfitting. In this study, we used the scikit-survival package that includes this method [29].

#### 2.3.4 Gradient boosting survival model (GBS)

GBS is an ensemble learning method that combines the predictions of multiple weak base learners to create a powerful overall model. Unlike random forest, GBS combines the base learners into a weighted ensemble model, giving extra weight to weak learners for correct predictions. In survival analysis, the loss function is derived from the Cox partial likelihood function and regression trees are used as base learners [30].

#### 2.3.5 Survival support vector machines (SVM)

SVM are supervised learning models initially used to maximize the margin between classes and to find the separating hyperplane that minimizes misclassification between classes. In our case SVMs are employed to learn a ranking based all possible pairs of patients rather than a binary classification. More specifically, the SVM is trained to assign patients with shorter survival a lower rank than those with longer survival [31]. The observations on the margin of the decision hyperplane are known as support vectors. SVM can manage non-linear relationships between features and survival data using the kernel trick. The kernel function transposes the input features into high-dimensional feature spaces where a linear ranking function functioned can be estimated.

### 2.4 Data pre-processing

To train the survival models, the input feature sets underwent various data pre-processing steps that included handling missing values, scaling numerical variables, and encoding categorical variables using one-hot encoding. For missing value imputation, we used Iterative Imputer with the Random Forest classifier as the imputation model. This strategy models each feature with missing values as a function of other features and uses the function to estimate values for imputation. As the features with missing values were categorical in nature, we opted for a Random Forest classifier. We fitted the imputer to the training dataset only and then applied it to transform the test dataset. Imputation was carried out in the outer loop of the nested cross-validation procedure, and a random seed was used to reproduce the same imputed values for all feature sets across repetitive trials. This helped to ensure the feature set comparability.

As the clinical features included predominantly categorical variables, we used one-hot encoding to transform them into numerical variables. We scaled continuous variables to a range of [0, 1] using max-min normalization to ensure that the same scales were used for all input variables.

### 2.5 Nested cross validation

We employed a nested, stratified 5-fold cross-validation to train and tune hyperparameters for the time-to-event models. The dataset was split into five outer folds, while balancing samples from different sources. In each outer fold, the pre-processed training set was further split into three inner folds for training and tuning hyper-parameters. We selected the model and hyperparameters with the highest concordance index across the inner folds to evaluate the outer test dataset. We repeated this procedure ten times for all survival models with all feature sets for reproducibility. We used random seeds as necessary.

### 2.6 Evaluation of the model prediction performance

#### 2.6.1 Concordance index

The Concordance Index (C-index) proposed by Uno et al [32] is a widely used performance metric to compare survival models applied to right-censored data. The C-index value is defined by the proportion of concordant predictions and outcomes of all comparable pairs. It defines the number of correctly ordered pairs by models. A risk score is attributed to each sample. Samples with a higher estimated risk score have a shorter actual survival time. Two samples are considered a comparable pair if they both experienced an event at two different times or if the one with a shorter observed survival time experienced an event, in which case the event-free subject “outlived” the other. Two samples are not comparable if they experienced events at the same time.

#### 2.6.2 Time-dependent area under the ROC curve (AUC)

When predicting survival, the patients disease status is not fixed and can change over time. Consequently, performance measures, such as specificity and sensitivity, become time-dependent measures. We considered an estimator of the cumulative/dynamic AUC at specific time points to evaluate our models [33]. [31]. The function considers comparable pairs of instances [i.e. one instance experienced an event before time *t* (*t*_*i*_ ≤ *t*) and the other at time *t*_*i*_ *> t*, and calculates the AUC at discrete time points. The time-dependent AUC for any specific survival time *t* can be calculated as follows:

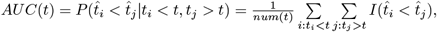

where *t* ∈ *T* is the set of all possible survival times, *t*_*i*_ and 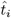 represent the observed time and the predicted value respectively, *num*(*t*) is the number of comparable pairs at time *t* and *I*(.) is the indicator function [34]. Then, we can then determine how well a model can distinguish subjects who fail (i.e. disease progression) before given time (*t*_*i*_ ≤ *t*) from subjects who fail after this time *t*_*i*_ *> t*.

#### 2.6.3 Time-Dependent Brier score

In survival analysis, the time-dependent Brier score (BS) is a crucial metric for assessing the model calibration over time because it takes into account dynamic changes in survival probabilities. It quantifies the concordance between predicted and observed outcomes at various time points, offering a comprehensive evaluation of model’s calibration [35]. We consider that for each patient, at *t* = 0, we have information on a vector **X** of patient specific covariates. This is used to predict a time-to-event variable *T*. Let us suppose that the complete data (*T*_*i*_, **X**_*i*_), *i* = 1, …, *n* is available for *n* patients. We also consider that *T* is censored and we observe 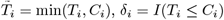, where *C*_*i*_ are the (hypothetical) times under observation. Under the common assumption of random censorship (*T* and **X** are independent on *C*) the empirical Brier score is defined as:

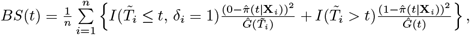

where 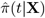 is the predicted probability of remaining event-free up to time point *t* given the feature vector **X** and *Ĝ*(*t*) is the probability of censoring weight, estimated by the Kaplan-Meier estimator.

Additionally, the integrated Brier score provides a concise measure of calibration across the entire time period:

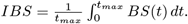

#### 2.6.4 Wilcoxon signed-rank test

We used the Wilcoxon signed-rank test to compare the performance of the survival models. This non-parametric test compares paired differences in the C-index, derived from the same sample, using a significance level of 0.05. We ensured reproducibility by using a consistent random state, resulting in the same training and test datasets in identical fold conditions for each model. Consequently, we could pair the C-indexes of two models. We then compared the performance of the top-performing models and feature sets. We also used a one-sided test, at 5% significance, to evaluate whether the median of the better-performing model was higher, effectively testing whether the highest-ranked model performance was significantly better.

### 2.7 Code availability

The open source code has been made available for the computational biology community: https://github.com/sarahlne/MelanomaMLProject

## 3 Results

### 3.1 Patient characteristics and genomic data pre-processing

The clinical features of the four patient cohorts did not show any significant imbalance (Table 1). Although the number of sequenced genes greatly varied among cohorts (from full exon sequencing to a list of 35 genes), the percentage of mutated genes in tumors before treatment was comparable among cohorts (Table 1). Only in the Catalanotti *et al*. cohort, the mean percentage of mutated genes per patient was higher, although the number of sequenced genes was intermediate compared with the other cohorts.

Melanoma is among the tumors with the highest mutational burden [36], making it difficult to distinguish between disease-driver and neutral mutations. To extract features that might be informative for predicting the response to treatment, we pre-processed the initial mutational data using three strategies and obtained eight different genomic datasets (Fig. 1 and Supplementary Table S1). The first strategy was to select genes with mutations that had a functional consequence (*Fathmm, Cscape* and *Cscape*^*High*^ datasets). The second strategy was to select genes involved in melanoma signaling pathways (*KEGG-Mel* and *VCELLS-Mel* datasets). The third strategy was to consider only the mutation rate in generic (*MR*^*KEGG*^ dataset) or melanoma (*MR*^*KEGG-Mel*^ and *MR*^*VCELLS-Mel*^ datasets) signaling pathways.

**Figure 1:**
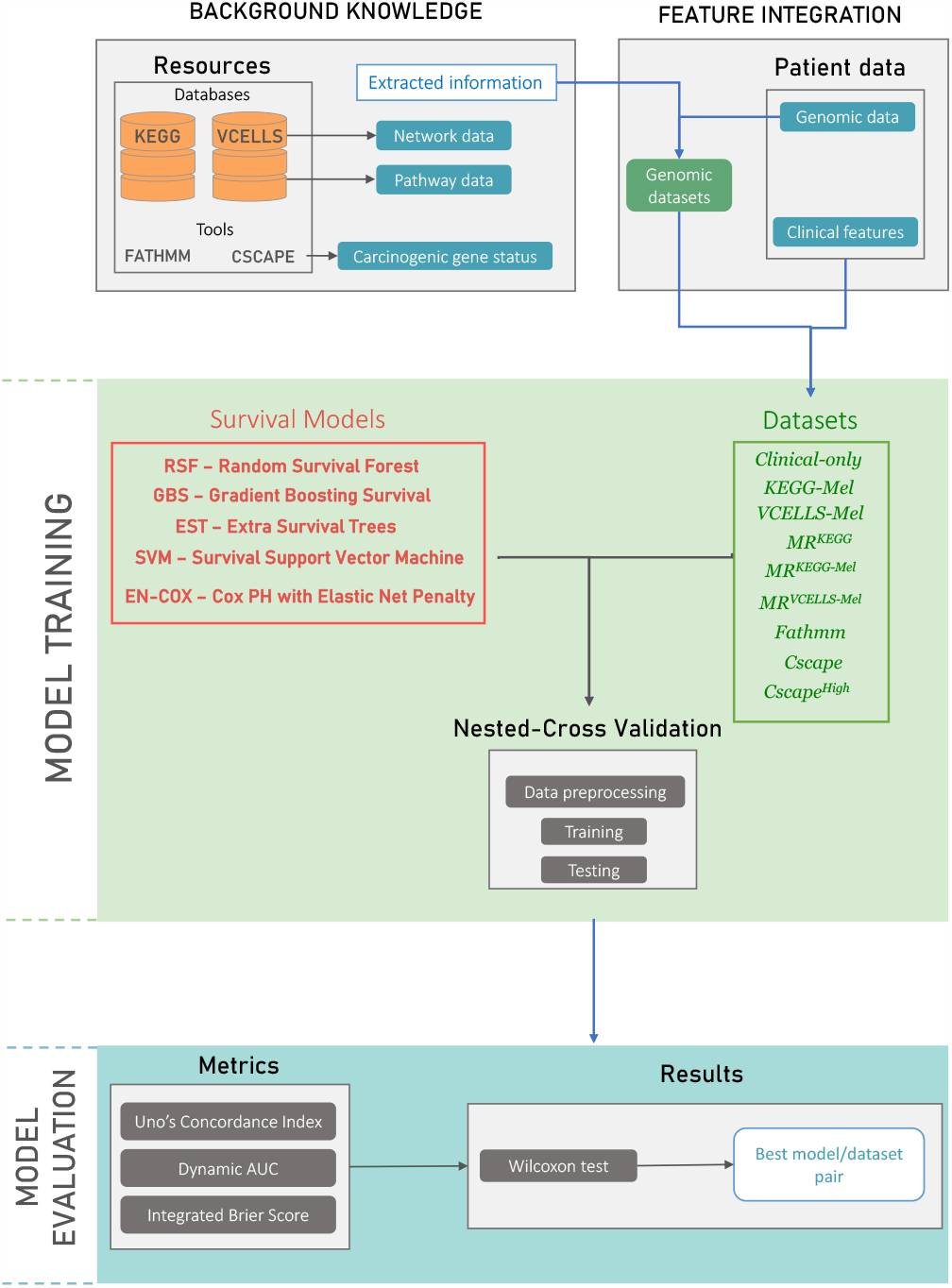
Overall scheme of information extraction, training and testing procedures. Background knowledge (e.g. melanoma network and signaling pathway data) were extracted from the KEGG and VCELLS databases. FATHMM and Cscape were used to retrieve putative cancer-promoting/driver mutation. These background knowledge was then used in the feature integration section to define features from the patients genomic data. Four survival analysis models were trained using the generated genomic datasets after data pre-processing through a 5-fold nested-cross validation process. For model evaluation, survival analysis metrics (i.e. Uno’s C-index, dynamic AUC and integrated Brier score) were used to assess the model performance. Finally, the Wilcoxon signed-rank test was used to identify the best performing algorithm/data set pair.

### 3.2 Evaluation of the model performance

To evaluate the value of baseline genomic data for predicting the response to MAPKi, we compared the results obtained using clinical features alone and supplemented with each of the eight genomic datasets (Supplementary Table S1). To train and test our models, we chose to work with the two patient cohorts with the highest number of sequenced genes (Catalanotti *et al*.; Van Allen *et al*.). As the prediction was constrained by few data and cases, we embedded the best overall response in the data used for the prediction. We then applied a panel of statistical and machine learning algorithms adapted to survival analysis to predict progression-free survival during treatment (Figure 1).

All models trained using the *Clinical-only* showed a good C-index (0.72 to 0.74), and the RSF and SVM algorithms displayed the best score (0.74) (Figure 2a). We observed a similar stability for the models trained using the *MR*^*KEGG-Mel*^ and *MR*^*VCELLS-Mel*^ datasets that contained only one additional variable compared with the *Clinical-only* dataset. Conversely, other models trained with clinical and genomic data and containing more variables had more fluctuating scores in function of the algorithm, for example between 0.63 and 0.74-0.75 for the *MR*^*KEGG*^ and *Cscape* datasets. This suggested that the addition of variables made the models more sensitive to the type of algorithm. Nevertheless, when we averaged the scores obtained with the different datasets, we observed that all algorithms displayed a C-index between 0.71 and 0.74.

**Figure 2:**
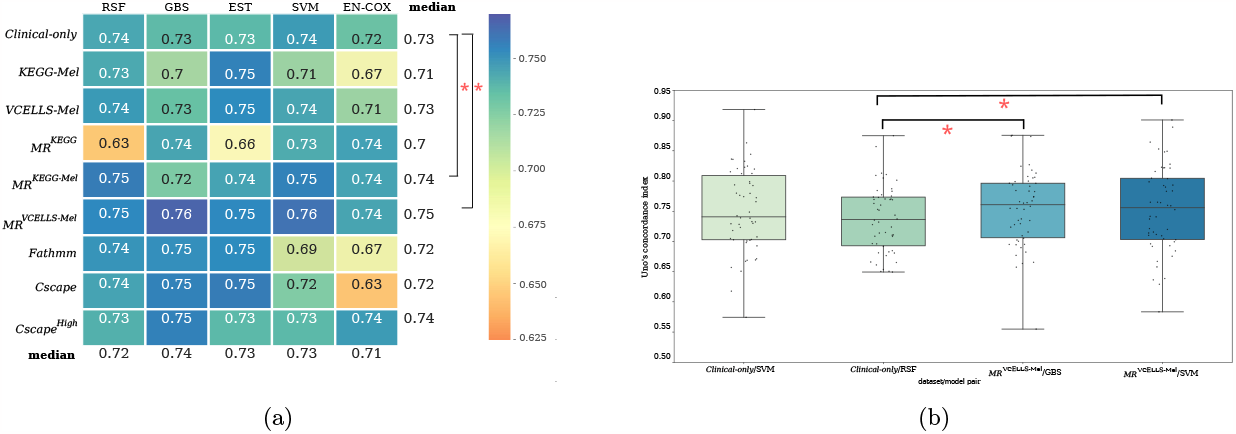
Prediction of the model performance using the concordance index (C-index). (a) Heatmap showing the performance of each dataset/algorithm pair evaluated using the Uno’s C-index. The median C-index was calculated though the training and testing procedure and is represented as a color gradient according to the distribution of the obtained values (0.63-0.76). The median performance values for each dataset with all algorithms and for each algorithm using all datasets are shown in the last row and last column, respectively. The Wilcoxon signed-rank test was used for pair comparison; *p <0.05 (versus the *Clinical-only* dataset). (b) Boxplot showing the C-index values for the best dataset/algorithm pairs: (*MR*^*VCELLS-Mel*^/GBS and SVM) compared with the best performing *Clinical-only* dataset/algorithm (RSF and SVM).

Among the models trained on clinical features supplemented with genomic data, those trained using the *MR*^*KEGG-Mel*^ and *MR*^*VCELLS-Mel*^ datasets had a significantly higher mean C-index than models trained using clinical features alone (Figure 2a). Specifically, the best dataset/algorithm pairs were *MR*^*VCELLS-Mel*^/GBS and *MR*^*VCELLS-Mel*^/SVM. Indeed, they displayed significantly higher C-index values than the *Clinical-only* /RSF OR SVM pair (Figure 2b). Additionally, the models trained using the *MR*^*VCELLS-Mel*^ dataset had a significantly lower integrated Brier score (i.e. better calibration) than models trained using clinical features alone (Supplementary Figure 1). They also performed better than models trained on clinical features alone at all time points, as indicated by the time-dependent AUC values (Supplementary Figure 2). This means that the addition of the mutation rate of genes implicated in melanoma signaling pathways improved the prediction obtained with clinical features alone. This effect was less important when the lists of mutated genes were added.

### 3.3 Validation of the predictive value of baseline genomic data

To validate the predictive value of our models, we chose to test them using data from the two patient cohorts not used for training (Blateau *et al*.; Louveau *et al*.). Genomic data from these two cohorts were generated by targeted sequencing of a limited number of cancer-related genes, carried out routinely for diagnostic purposes at these two hospital centers. To take into account the bias due to the lower number of genes sequenced in these two cohorts, we only used the genomic datasets with the mutation rate. These were also the ones that produced the best scores in the training step. The C-index score of the *MR*^*KEGG*^/GBS pair was higher than that of the Clinical-only/SVM pair (0.78 vs 0.76) *Clinical-only* /SVM pair (Figure 3). This result validated the improved prediction of the response to MAPKi by adding baseline genomic data.

**Figure 3:**
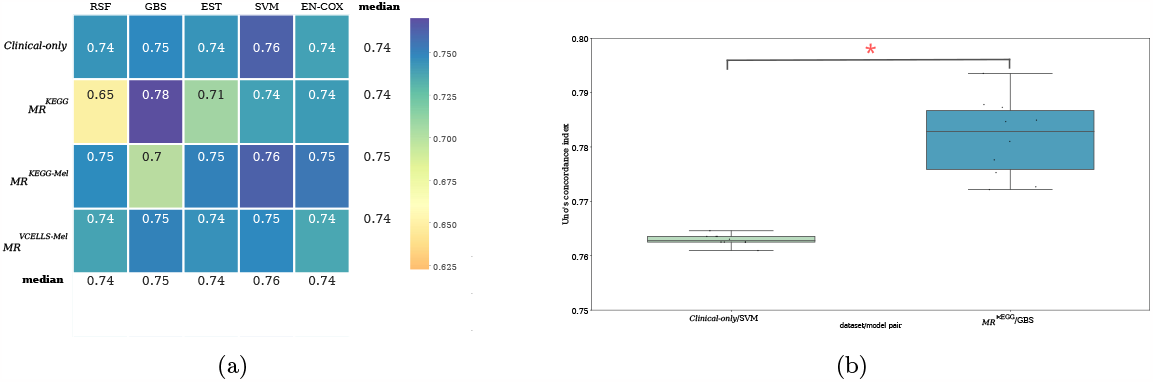
Prediction performance in the two validation cohorts using the C-index. (a) Heatmap representation showing the performance of each dataset/algorithm pair using the Uno’s C-index. Algorithms were trained using the training cohorts (Catalanotti *et al*.; Van Allen *et al*.) and tested using data from the other two cohorts (Blateau *et al*.; Louveau *et al*.) The median C-index values are represented as a color gradient according to their distribution (0.65-0.78). The median performance values for each dataset using all algorithms and for each algorithm using all datasets are shown in the last row and last column, respectively. (b) Boxplot showing the C-index values for the best dataset/algorithm pairs: *Clinical-only* /SVM and *MR*^*KEGG*^/GBS. *pvalue<0.05.

### 3.4 Assessment of the improved prediction in clinical context

To assess the improved prediction of our models in a clinical context, we built two scenarios and tested them by censoring data in line with each scenario. In the first “duration of clinical benefit” scenario, patients would start MAPKi treatment, and those with disease progression would stop treatment in the first few months. In this scenario, the question was to predict the duration of clinical benefit for the remaining patients. For this scenario, we censored patients with progressive disease as best overall response. In the second scenario “progression before 12 months”, the question was to predict before treatment initiation, which patients would progress during treatment before 12 months. For this scenario, we censored progression-free survival data at 12 months.

To train and test our models, we chose to work with the two patient cohorts that had the highest number of sequenced genes (Catalanotti *et al* and Van Allen *et al*) and with the genomic datasets that included the mutation rate. Overall, the C-index values were lower than those obtained during the evaluation and validation steps, certainly due to data censoring (Figure 4). Nevertheless, we still obtained a trend of improved prediction when we added the baseline genomic data. For the “duration of clinical benefit” scenario, the best score was obtained by the *MR*/SVM pair (C-index = 0.7) compared with the *Clinical-only* /SVM pair (C-index = 0.69) (Figure 4a). For the “progression before 12 months” scenario, the median C-index values of the *MR*^*VCELLS-Mel*^/RSF pair and *Clinical-only* /RSF pair were 0.64 and 0.61, respectively (Figure 4b). These results highlight the value of baseline genomic data for predicting the duration of the response to MAPKi in two clinical setting scenarios.

**Figure 4:**
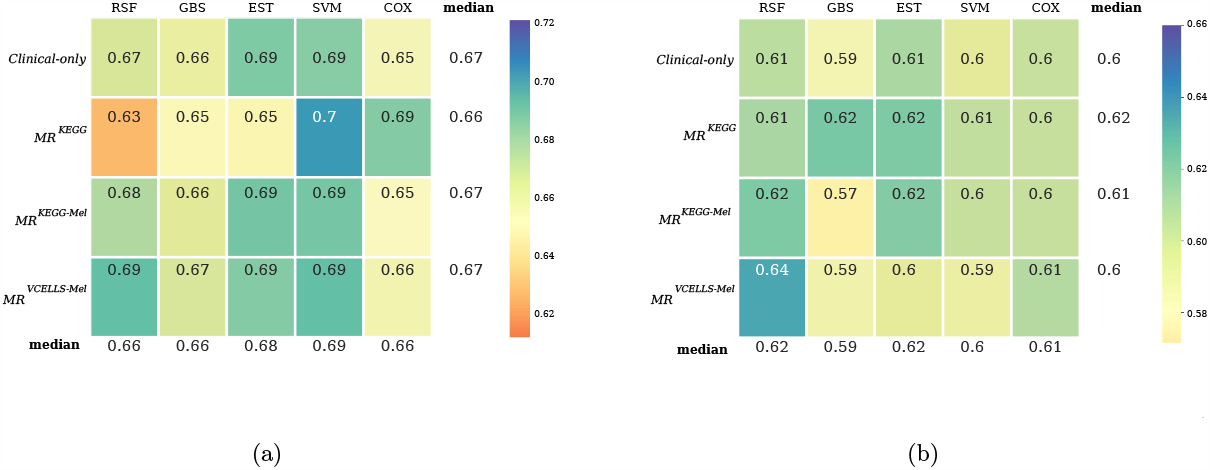
Prediction performance in clinical scenarios. Heatmap showing the performance of each dataset/algorithm pair evaluated using the Uno’s C-index. (a) “Duration of clinical benefit” scenario and (b) “Progression before 12 month” scenario. The median C-index was calculated through the training and testing procedures and is represented as a color gradient according to the distribution of the obtained values. The median values for each dataset using all algorithms and for each algorithm using all datasets are shown in the last row and last column, respectively.

## 4 Discussion

In this study, we tested whether baseline genomic data could improve the prediction of the response to MAPKi in patients with advanced BRAF^V600^-mutated melanoma. To the best of our knowledge, machine learning approaches had never been used for this purpose. We used several algorithms in combination with different strategies for pre-processing tumor mutation data. Our results show that baseline (before treatment) tumor mutation data contain information that can be used to better predict progression-free survival.

This observation was not obvious because progression-free survival is influenced by primary and also acquired resistance. Although primary resistance is mainly linked to the initial tumor cell status and their microenvironment, to date there is still no biomarker to identify patients with BRAF^V600^-mutated melanoma who will not respond to MAPKi. Acquired resistance involves the adaptation of tumor cells through genetic or non-genetic mechanisms [37, 38]. A single-cell study showed that tumor heterogeneity and melanoma cell plasticity influence tumor behavior and evolution under MAPKi pressure [39]. Therefore, it would seem counter-intuitive to detect information relevant for predicting the response duration using pre-treatment bulk genomic sequencing data. The mechanistic relationships between the genetic context before treatment and the tumor response to treatment, including plasticity phenomena, are subjects for future research.

We acknowledge the constraints and limitations inherent in our study. First of all, our sample size was small, and we used data from different cohorts from different centers. Indeed, we grouped together patients treated with a BRAF inhibitor and patients treated with a combination of BRAF and MEK inhibitors. Despite this diversity, we found that baseline genomic data improved the treatment response prediction. If the existing data were more easily accessible, we could work with more homogeneous patient cohorts. This would allow us to determine whether this observation is specific to the molecule administered. Second, the majority of sequencing data were only partial, and therefore, we probably missed gene alterations relevant to the prediction of the response to MAPKi. However, our results demonstrated that partial sequencing, which is routinely used in clinical practice, already provides potentially valuable information for predicting the response to MAPKi. A future analysis using whole exome sequencing data could allow determining the minimal and optimal gene set for prediction.

Our results demonstrate the predictive potential of baseline genomic data, and the value of integrating them in the development of predictive models for the response to MAPKi in the context of advanced melanoma. They also show that better exploiting the sequencing data already available at cancer centers will open new avenues for therapeutic decision-making.

## Data Availability

All data produced in the present study are available upon reasonable request to the authors.

## Acknowledgements

This work has been supported by the INCa-Cancéropôle GSO (Programme Emergence Intelligence Artificielle N°2022-IA20 and Programme Emergence N°2018-E1), the Fondation ARC pour la recherche sur le cancer (Projets ARC 2022 PJA2 N°ARCPJA2022060005107), and the Ligue nationale contre le cancer (Comité du Gard JPB/GA/MV/39-2019). SD is supported by the Agence nationale pour la recherche (Intelligence Artificielle en Santé et Environnement – AXIAUM N°ANR-20-THIA-0005-01) and the University of Montpellier/Ecole Doctorale CBS2.

## Supplementary data

**Supplementary Table S1:** Description of the dataset composition.

| Dataset | Description | Feature composition | Total # feature |
| --- | --- | --- | --- |
| *Clinical-only* | Clinical features | 8 | 8 |
| *KEGG-Mel* | Clinical features + KEGG melanoma network data | 8+70 | 78 |
| *VCELLS-Mel* | Clinical features + VCELLs melanoma network data | 8+119 | 127 |
| *MR<sup>KEGG</sup>* | Clinical features + All pathway probabilities (KEGG+VCELLs) | 8+332 | 340 |
| *MR<sup>KEGG-Mel</sup>* | Clinical features KEGG melanoma pathway probabilities | 8+1 | 9 |
| *MR<sup>VCELLS-Mel</sup>* | Clinical features + VCells melanoma pathway probabilities | 8+1 | 9 |
| *FATHMM* | Clinical features + fathmm data (5% threshold) | 8+39 | 47 |
| *Cscape* | Clinical features + Cscape data | 8+53 | 61 |
| *Cscape<sup>High</sup>* | Clinical features + Cscape data (only high confidence genes) | 8+171 | 179 |

**Supplementary Figure 1:**
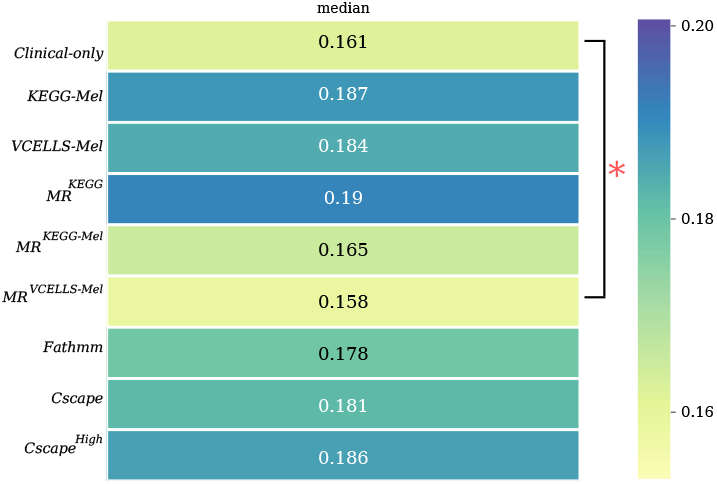
Integrated Brier scores. The median integrated Brier score was calculated from the Brier scores obtained through the training and testing procedures. Values are represented as a color gradient according to the distribution of the obtained values (0.161-0.19). The Wilcoxon signed-rank test was used to feature dataset/algorithm pair comparison; *p-value<0.05 versus the *Clinical-only* dataset

**Supplementary Figure 2:**
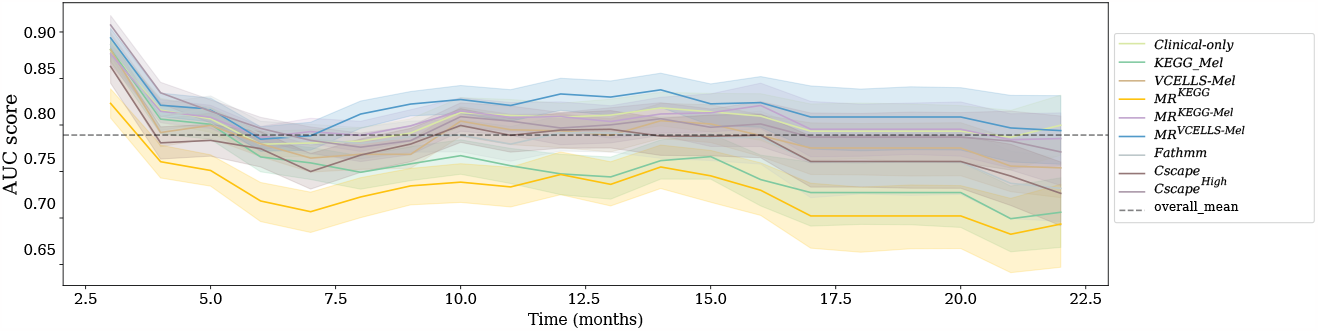
Evaluation of model predictions over time (Time-dependent AUC) Comparison of the performance of the feature datasets for all algorithms over 22 months. Each line refers to the mean AUC value of that feature datasets at each time point with their associated standard deviation.

## Supplementary materials and methods

### Survival analysis algorithms

#### Elastic net penalized Cox regression model

Penalized models introduce regularization by adding penalty terms to the model equation. The Elastic Net penalty regularization method is particularly useful in scenarios with more features than observations. It combines the strengths of lasso (*l*_1_) and ridge (*l*_2_) penalties, by performing feature selection like lasso and by shrinking correlated features together as done in ridge regression [27]. In the Cox regression model, regularization is added to the log partial likelihood, resulting in the following equation:

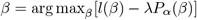

where *l*(*β*) is the log partial likelihood, *λ* ≥ 0 the regularization parameter and *P*_*α*_(*β*) a penalty form. In the elastic net penalized Cox regression model, the penalty term is expressed as:

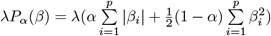

where *p* is the number of variables and *α* ∈ (0, 1] is the additional parameter that controls the amount of shrinkage that comes from the *l*_1_ and *l*_2_ penalties. Setting *α* = 1 corresponds to the lasso penalty.

#### Random survival forest (RSF)

RSF is an ensemble method used for survival analysis that uses survival trees as its base model. Unlike the decision trees used in random forests (described by Breiman), RSF employs survival trees and averages the ensemble cumulative hazard function of each tree for prediction [28]. RSF is based on the bagging method in which random samples are repeatedly drawn from the input data to reduce the base model variance and avoid overfitting. However, an extra level of randomness is added to RSF while growing the trees. This random subset of features is used for splitting a node instead of using all features. This reduces the correlation among trees in the forest and improves the prediction performance. Each node of the tree is split using the random subset of features to maximize the dissimilarity between child-nodes. The non-parametric Nelson-Aalen estimator is used to estimate the ensemble cumulative hazard function by taking the average of all cumulative hazard functions of each survival tree [30, 34, 40].

#### Extra survival trees (EST)

EST, also known as extremely random survival forest, is a meta-estimator developed as an extension of the random survival forest method. This method fits several randomized survival trees, called extra-trees, and like RSF, uses averaging to improve the predictive accuracy and to control overfitting. The extra level of randomness in extra survival trees comes from how splits are computed, which is different from the method used in RSF. Instead of taking the most discriminative thresholds at each node, thresholds for each feature are drawn randomly, and the best of these randomly generated thresholds is used for splitting the node. This approach tends to improve the model generalization by reducing the overall model bias and variance. The log-rank statistic is used as a splitting criterion for extra survival trees, like in RSF. The scikit-survival package includes this method, and it was used in the current study in addition to other models [29].

#### Gradient boosting survival (GBS)

Gradient boosting is an ensemble learning method that combines the predictions of multiple weak base learners to create a powerful overall model. Unlike random forest, gradient boosting combines the base learners into a weighted ensemble model, giving extra weight to weak learners for correct predictions. This algorithm updates base learners in an additive manner. The addition of each base learner boosts the overall model. Gradient boosting optimizes the loss function using a gradient descent algorithm in which the steepest gradient step is taken at each boosting step. The model sequentially minimizes the residual error and improves prediction performance ([34, 41].

In survival analyses, the loss function is derived from the Cox partial likelihood function, and regression trees are used as base learners [30]. Specifically, the linear model in the partial likelihood function of the Cox model is replaced by an additive model, and the objective of the loss function is to maximize the log partial likelihood. Gradient boosting survival models are constructed sequentially in a greedy stagewise fashion.

#### Survival support vector machines (SVM)

SVM are supervised learning models initially used to maximize the margin between classes and find the separating hyperplane that minimizes misclassification between classes. The observations on the margin of the hyperplane are known as support vectors. SVM can manage non-linear relationships between features and survival data using the kernel trick. The kernel function transposes the input features into high-dimensional feature spaces where a linear survival function can be estimated.

SVM were initially proposed for solving binary classification problems and were then extended to other problems, such as regression, clustering, ranking, and finally censored targets in survival analysis [31]. Survival analysis in with SVM can be tackled in two different ways: a) by ranking samples according to their survival times, and b) by using a regression approach in order to find a function that estimates the observed survival times as continuous outcomes.

